# Prevention of adrenal crisis: cortisol responses to major stress compared to stress dose hydrocortisone delivery in adrenal insufficiency

**DOI:** 10.1101/2020.02.08.20021246

**Authors:** Alessandro Prete, Angela E Taylor, Irina Bancos, David J Smith, Mark A Foster, Sibylle Kohler, Violet Fazal-Sanderson, John Komninos, Donna M O’Neil, Dimitra A Vassiliadi, Christopher J Mowatt, Radu Mihai, Joanne L Fallowfield, Djillali Annane, Janet M Lord, Brian G Keevil, John AH Wass, Niki Karavitaki, Wiebke Arlt

## Abstract

**Context:** Patients with adrenal insufficiency require increased hydrocortisone cover during major stress to avoid life-threatening adrenal crisis. However, current treatment recommendations are not evidence-based.

**Objective:** To identify the most appropriate mode of hydrocortisone delivery in patients with adrenal insufficiency exposed to major stress.

**Design and Participants:** Cross-sectional study: 122 unstressed healthy subjects and 288 subjects exposed to different stressors (major trauma [N=83], sepsis [N=100], and combat stress [N=105]). Longitudinal study: 22 patients with preserved adrenal function undergoing elective surgery. Pharmacokinetic study: 10 patients with primary adrenal insufficiency undergoing administration of 200mg hydrocortisone over 24 hours in four different delivery modes (continuous intravenous infusion; six-hourly oral, intramuscular or intravenous bolus administration).

**Main Outcome Measure:** We measured total serum cortisol and cortisone, free serum cortisol and urinary glucocorticoid metabolite excretion by mass spectrometry. Linear pharmacokinetic modelling was used to determine the most appropriate mode and dose of hydrocortisone administration in patients with adrenal insufficiency exposed to major stress.

**Results:** Serum cortisol was increased in all stress conditions, with the highest values observed in surgery and sepsis. Continuous intravenous hydrocortisone was the only administration mode persistently achieving median cortisol concentrations in the range observed during major stress. Linear pharmacokinetic modelling identified continuous intravenous infusion of 200mg hydrocortisone over 24 hours, preceded by an initial bolus of 50-100mg hydrocortisone, as best suited for maintaining cortisol concentrations in the required range.

**Conclusions:** Continuous intravenous hydrocortisone infusion should be favored over intermittent bolus administration in the prevention and treatment of adrenal crisis during major stress.

## INTRODUCTION

The activation of the hypothalamic-pituitary-adrenal axis in response to stressful stimuli elicits increased glucocorticoid output, aiming at restoring homeostasis. Cortisol is the major glucocorticoid produced by the human adrenal glands and is a key component of the physiological stress response (1).

Adrenal insufficiency is caused by failure of the adrenal cortex to produce cortisol, which can be caused by loss of function of the adrenal itself or its hypothalamic-pituitary regulatory center or, most commonly, long-term exogenous glucocorticoid treatment for other conditions. Patients with adrenal insufficiency are unable to produce adequate amounts of cortisol in response to stress and, therefore, require increased hydrocortisone replacement doses to avoid life-threatening adrenal crisis during surgery, trauma or severe infection (2-4). Prevention of adrenal crisis is challenging (5,6), and studies investigating the optimal dose and mode of steroid cover during major stress are lacking. Currently, administered hydrocortisone doses are chosen empirically rather than based on evidence. There is considerable variability in recommended administration modes, total doses, and dosing intervals (7). The lack of evidence-based recommendations for dose and mode of glucocorticoid replacement in major stress sends a confusing message to healthcare staff, which regularly exposes patients to harm (8).

This study was designed to determine the most appropriate hydrocortisone dose and delivery mode for patients with adrenal insufficiency during major stress. We employed tandem mass spectrometry to measure glucocorticoid concentrations in subjects with preserved adrenal function exposed to various conditions of stress, and compared them to concentrations achieved after administration of stress dose hydrocortisone by a range of currently used delivery modes in patients with adrenal insufficiency.

## MATERIALS AND METHODS

### Study design, participants, and procedures

Three clinical studies were undertaken (Figure 1), with patient demographics and outcome measures summarized in Table 1.

**Table 1.**
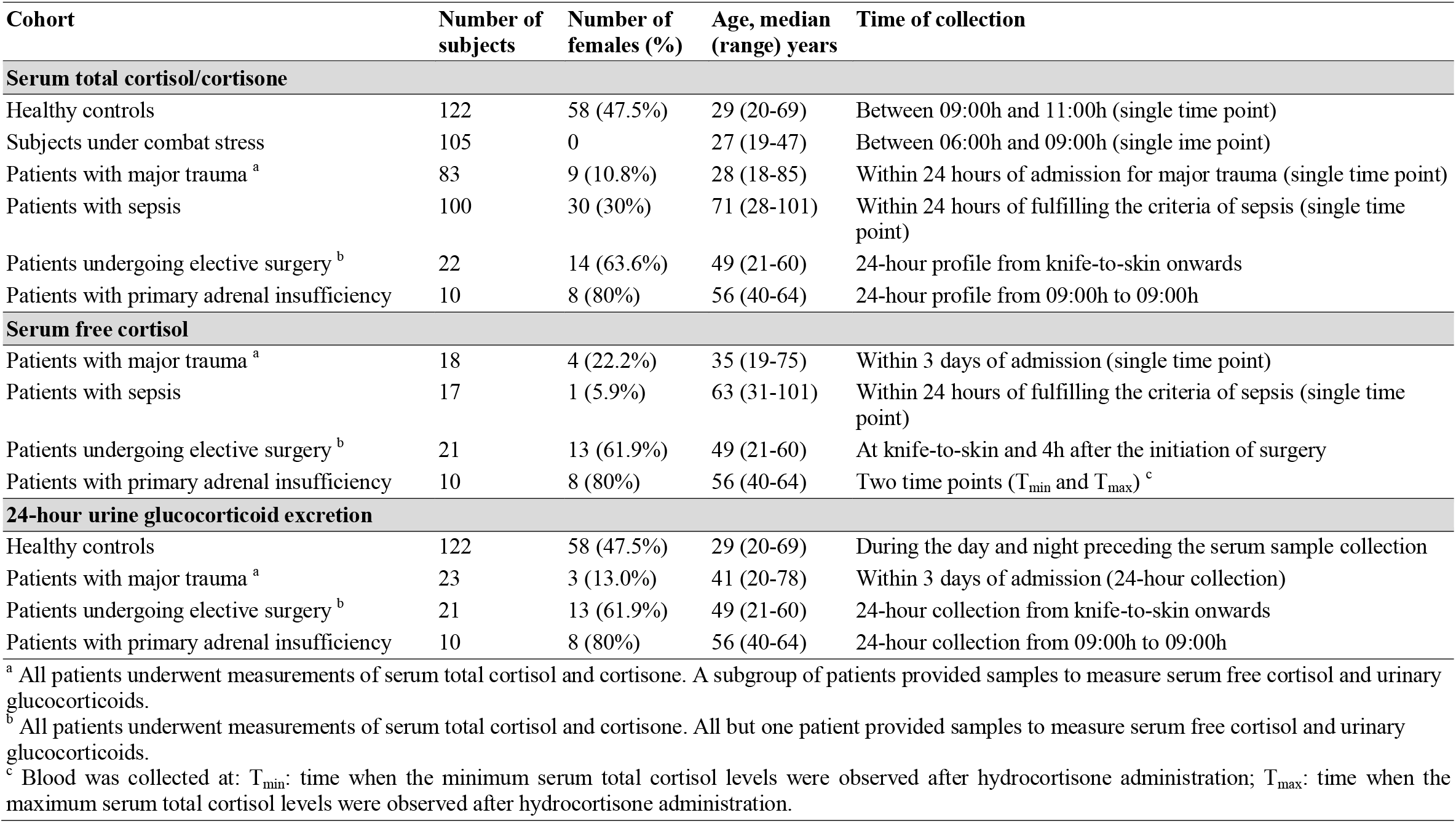
Clinical characteristics of study participants and sampling regimen.

**Figure 1.**
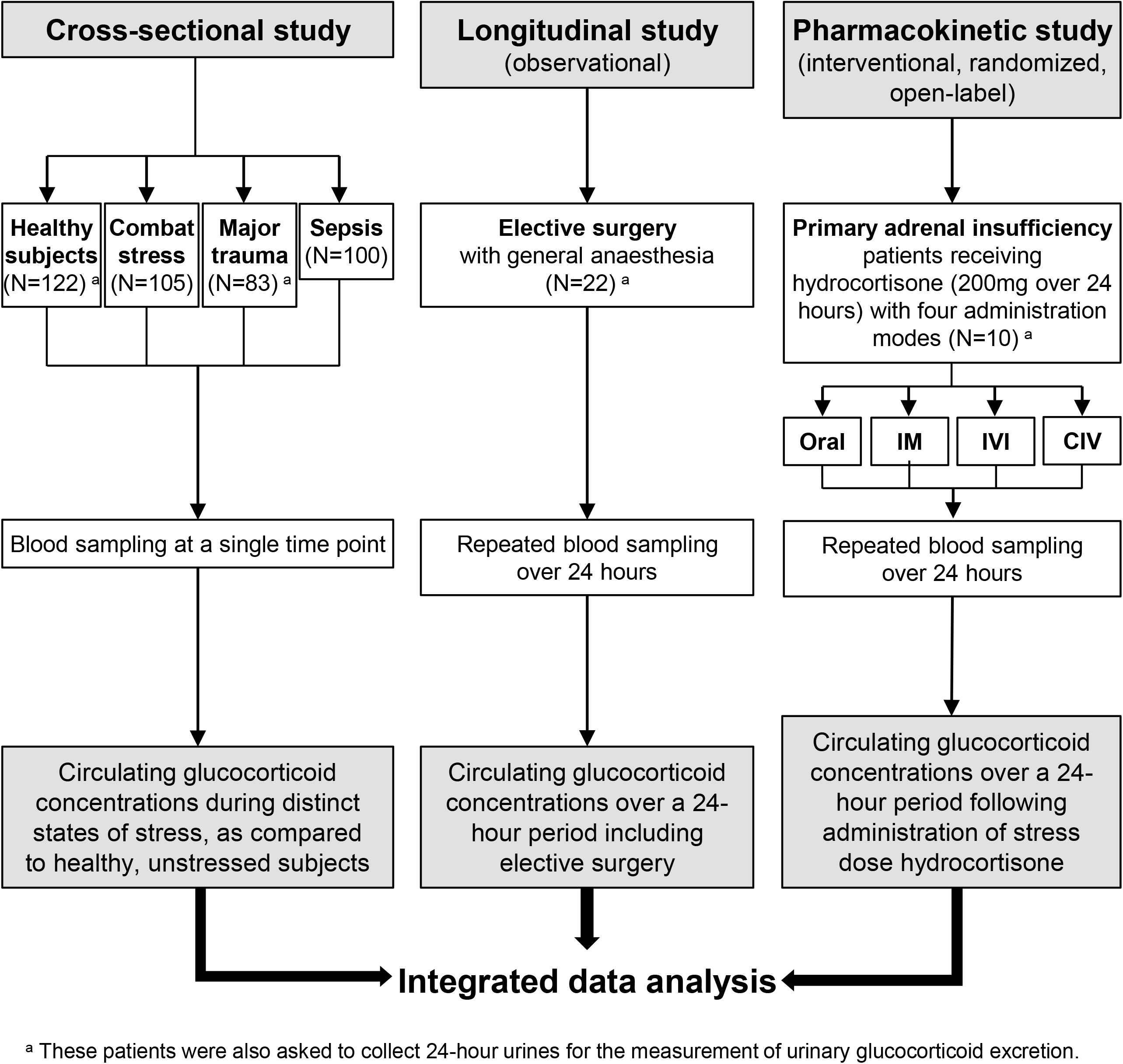
Summary of the studies performed. Assessment of the circulating and urinary glucocorticoid concentrations in response to different stress conditions and to stress dose hydrocortisone administration. Abbreviations: IM, intramuscular injection; IVI, intravenous injection; CIV, continuous intravenous infusion.

First, in a cross-sectional study, we measured circulating glucocorticoid concentrations in 122 healthy, non-stressed controls and 288 subjects with distinct and defined states of stress at the time of blood sampling. These conditions of stress included: 105 otherwise healthy subjects under combat stress (blood samples taken within four weeks of their deployment to the Afghanistan conflict) (9); 83 prospectively recruited subjects with acute major trauma (estimated new injury severity score [NISS] (10) >15; blood samples taken within 24 hours of acute injury, excluding brain injury); 100 consecutively recruited patients with sepsis (blood samples collected within 24 hours of fulfilling the criteria for sepsis (11) in the Intensive Care Unit setting). At the time of sampling, none of the subjects had an established diagnosis of adrenal insufficiency or were receiving treatment with glucocorticoids or other medications with major impact on steroid synthesis or metabolism.

Second, we prospectively recruited 22 patients with normal adrenal function who underwent repeated longitudinal serum sample collection over a 24-hour period whilst undergoing elective surgery with general anesthesia (Suppl. Table 1). Blood samples were drawn at the following time points: 0 (=knife-to-skin, KTS), 0.5, 1, 2, 3, 4, 5, 6, 12, and 24 hours.

Third, we undertook a randomized, open-label study in 10 patients with an established diagnosis of primary adrenal insufficiency and on stable steroid replacement therapy for at least six months (Suppl. Table 2). All patients attended the Clinical Research Facility for a 24-hour study period on four occasions separated by at least one week. On each study day, they were admitted at 08:00h after an overnight fast and last intake of their regular steroid replacement at 12:00h the preceding day; standardized meals were served at 10:00h, 14:00h, and 18:00h. On each of the study days, subjects received 200mg hydrocortisone over 24 hours administered by one of four different administration modes: oral tablets (ORAL; 50mg at 09:00h, 15:00h, 21:00h, and 03:00h); intramuscular bolus injection (IM; 50mg at 09:00h, 15:00h, 21:00h, and 03:00h); intravenous bolus injection (IVI; 50mg at 09:00h, 15:00h, 21:00h, and 03:00h); continuous intravenous infusion (CIV) of 200mg hydrocortisone over 24 hours (diluted in 50 ml glucose 5% and administered *via* perfusor at a rate of 4 ml/h). The four different administration modes were administered to each patient in random order (Suppl. Table 2). Blood sampling was carried out every 30 min from 9:00h-11:00h, 15:00h-17:00h, 21:00h-23:00h, and 03:00h-05:00h and otherwise in hourly intervals throughout the 24-hour study period.

### Ethics approval

All study participants provided written informed consent prior to inclusion and all study procedures underwent Ethics Committee approval prior to recruitment (combat stress: MOD REC 116/Gen/10; major trauma: NRES Committee South West – Frenchay 11/SW/0177; sepsis: Comité de Protection des Personnes de Saint-Germain-en-Laye – COITTSS trial NCT00320099; elective surgery and adrenal insufficiency: South Birmingham REC Ref 07/H1207/22).

### Glucocorticoid measurements

Serum concentrations of total cortisol and its inactive metabolite cortisone were measured by liquid chromatography-tandem mass spectrometry (LC-MS/MS) as previously described (12). For measurement of serum free cortisol concentrations, the unbound cortisol fraction was separated by temperature-controlled ultrafiltration, centrifuged in preconditioned ultrafiltration devices and then measured with LC-MS/MS, as previously described (13). Measurement of 24-hour urinary glucocorticoid excretion was carried out by gas chromatography/mass spectrometry, as previously described (14). For further details on the mass spectrometry analysis, see Suppl. Methods.

### Statistical analysis

Medians with 5^th^ to 95^th^ centile ranges and interquartile ranges were calculated for continuous variables. The area under the concentration-time curve (AUC) was calculated by means of trapezoidal integration. Serum cortisol concentrations between the various groups were compared by Kruskal-Wallis and Mann-Whitney U tests. The level of significance was set at *p*<0.05. Statistical analyses were performed by SPSS 178 21.0 for Windows (SPSS, Inc., Chicago, IL, USA) and MATLAB (Mathworks, Natick MA, USA).

### Pharmacokinetic modelling analysis

The serum cortisol time course response *c(t)* was modelled relative to intravenous hydrocortisone via linear pharmacokinetics, 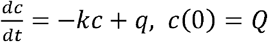, where *k* is clearance rate,*Q* is initial response (representing intravenous bolus [IVI] delivery), and *q* is rate of continuous intravenous (CIV) delivery of hydrocortisone. This model has the exact solution, 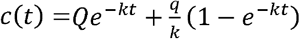. IVI 50mg data over 6-12 hours was used to fit the parameters *k* and *Q* (with *q* = 0) using a mixed effects model implemented in MATLAB (Mathworks, Natick MA, USA) and the function *nlmefit*. This approach enabled the estimation of population average (*fixed effects*) and between-patient heterogeneity (*random effects*). Responses to other modes of administration were predicted by plotting model solutions with appropriately modified parameters 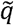 and 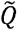 ; for example IVI 100mg was modelled by taking 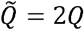 and 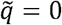; CIV 200mg/24h was modelled by taking 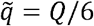 and 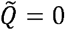.

## RESULTS

### Glucocorticoid concentrations in different conditions of stress

Serum total cortisol concentrations were highest and most variable in patients with sepsis, followed by patients undergoing elective surgery with general anesthesia, patients with combat stress, and patients with acute major trauma (Figure 2a). Pairwise comparisons showed significant differences between unstressed controls vs. all stressed groups, except for patients with major trauma.

**Figure 2.**
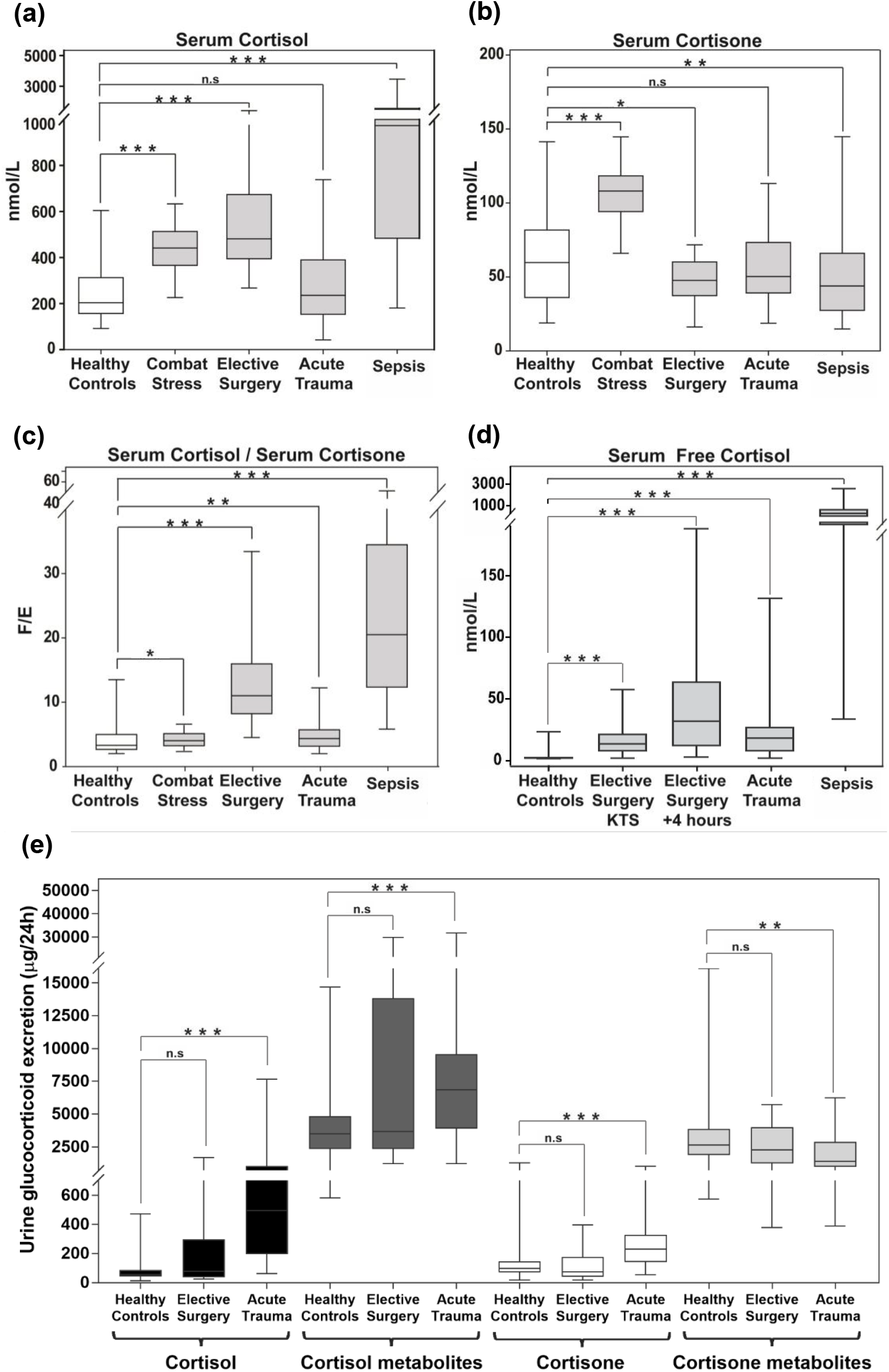
Circulating glucocorticoids during major stress. Serum concentrations of (a) total cortisol (nmol/L), (b) total cortisone (nmol/L), and (c) cortisol (F)/cortisone (E) ratio in healthy controls (N=122), during combat stress (N=105), during elective surgery (N=22), after major trauma (N=83), and during sepsis (N=100). In the patients undergoing elective surgery, the maximum serum cortisol levels (and corresponding serum cortisone levels) were used for the calculations. Panel d reports serum free cortisol concentrations in nmol/L in healthy controls (N=11), during elective surgery at knife-to-skin (KTS, N=21) and 4 hours after start of operation (N=21), after major trauma (N=18), and during sepsis (N=17). Panel e reports the 24-hour urinary excretion of cortisol, cortisol metabolites, cortisone, and cortisone metabolites in healthy controls (N=122), following elective surgery (N=21), and after major trauma (N=23). Boxes show median and interquartile range, whiskers are 5th to 95th percentile. Symbols: n.s., *p*>0.05; *, *p*≤0.05; **, *p*≤0.01; ***, *p*≤0.001.

When analyzing the inactive cortisol metabolite cortisone, pairwise comparisons to levels observed in unstressed controls showed significantly higher serum cortisone in combat stress, whilst circulating cortisone was significantly lower in elective surgery and sepsis patients; serum cortisone concentrations in patients after major trauma did not differ from unstressed controls (Figure 2b). The serum cortisol/cortisone ratio showed a significant increase, favoring active cortisol in all stress conditions, with the highest increase in sepsis (Figure 2c).

Free serum cortisol concentrations were higher than in unstressed controls in all stressed groups, with the highest concentrations observed in sepsis (Figure 2d).

24-hour urinary excretion of cortisol, cortisone and their major metabolites was significantly increased in major trauma, whilst glucocorticoid excretion in patients undergoing elective surgery did not significantly differ from unstressed controls (Figure 2e).

### Glucocorticoid dynamics during elective surgery

We separately analyzed circulating glucocorticoid concentrations in patients undergoing surgeries of short duration (median duration 60 min, range 25-85 min; n=11) from those who underwent longer-lasting surgery (median duration 175 min, range 100-295 min; n=11). In both groups, serum cortisol decreased within an hour of induction of anesthesia, followed by a gradual increase. In the group with shorter surgery, maximum serum cortisol concentrations (C_max_) were observed after a median of 3 hours post-KTS, while in the group with longer duration surgery, C_max_ were observed after a median of 5 hours post-KTS (Table 2 and Suppl. Fig. 1), i.e. during the wake-up phase after general anesthesia.

**Table 2.**
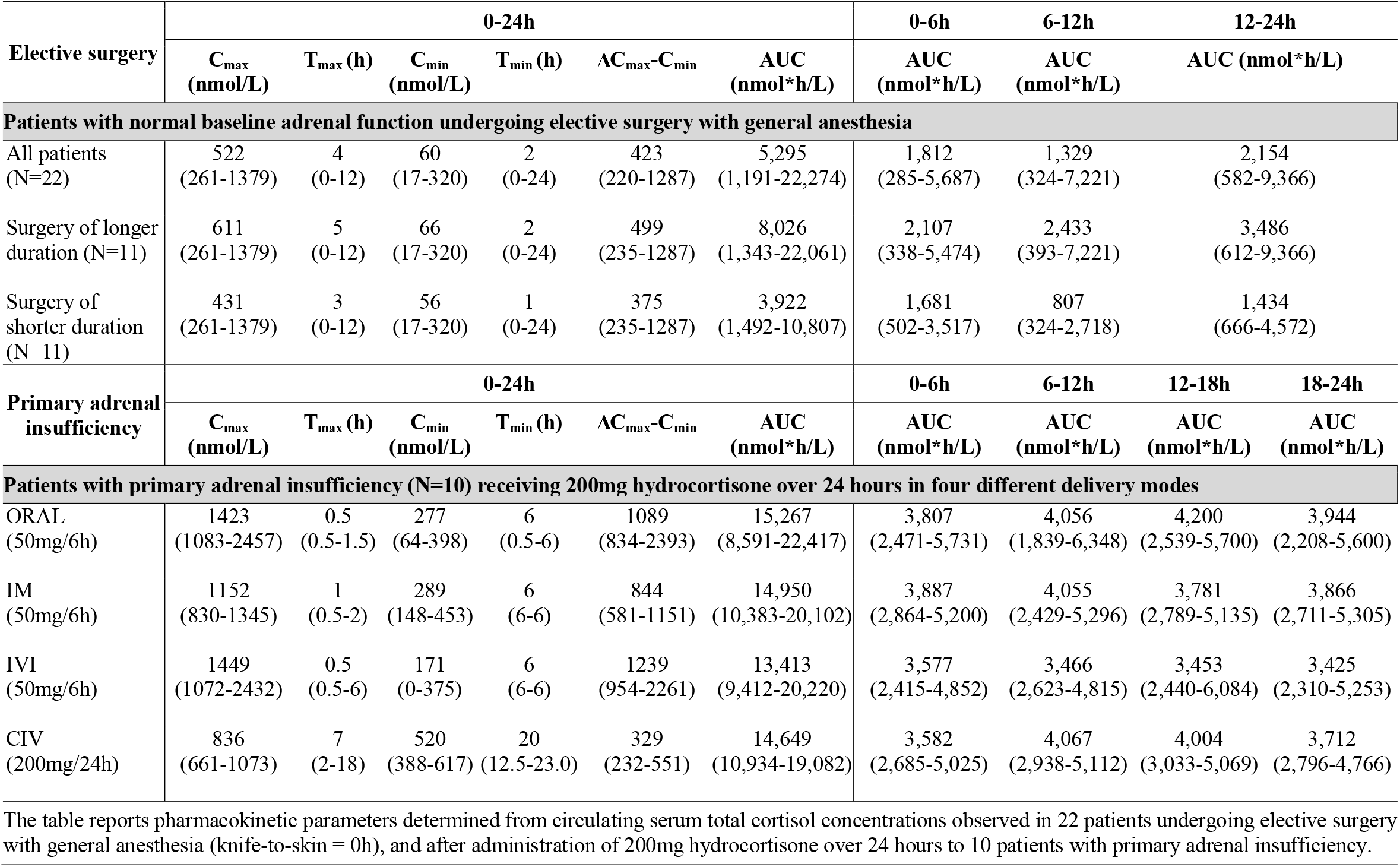
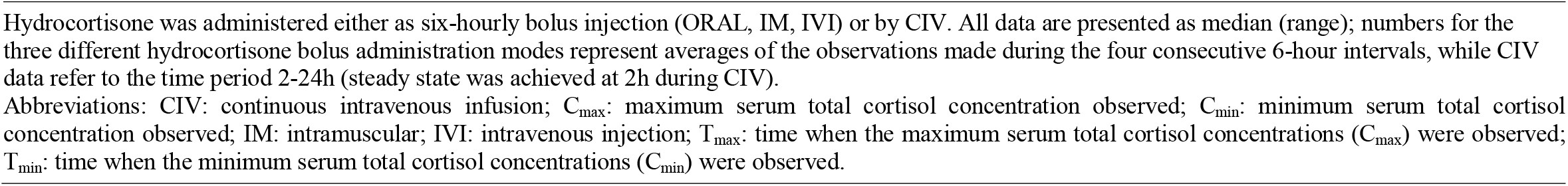
Pharmacokinetic parameters of serum total cortisol concentrations observed during elective surgery in patients with preserved adrenal function (N=22) and after hydrocortisone administration via four different modes in patients with primary adrenal insufficiency (N=10).

| Elective surgery | 0-24h |  |  |  |  |  | 0-6h | 6-12h | 12-24h |  |
| --- | --- | --- | --- | --- | --- | --- | --- | --- | --- | --- |
|  | C<sub>max</sub><br>(nmol/L) | T<sub>max</sub> (h) | C<sub>min</sub><br>(nmol/L) | T<sub>min</sub> (h) | ΔC<sub>max</sub>-C<sub>min</sub> | AUC<br>(nmol*h/L) | AUC<br>(nmol*h/L) | AUC<br>(nmol*h/L) | AUC (nmol*h/L) |  |
| Patients with normal baseline adrenal function undergoing elective surgery with general anesthesia |  |  |  |  |  |  |  |  |  |  |
| All patients<br>(N=22) | 522<br>(261-1379) | 4<br>(0-12) | 60<br>(17-320) | 2<br>(0-24) | 423<br>(220-1287) | 5,295<br>(1,191-22,274) | 1,812<br>(285-5,687) | 1,329<br>(324-7,221) | 2,154<br>(582-9,366) |  |
| Surgery of longer<br>duration (N=11) | 611<br>(261-1379) | 5<br>(0-12) | 66<br>(17-320) | 2<br>(0-24) | 499<br>(235-1287) | 8,026<br>(1,343-22,061) | 2,107<br>(338-5,474) | 2,433<br>(393-7,221) | 3,486<br>(612-9,366) |  |
| Surgery of<br>shorter duration<br>(N=11) | 431<br>(261-1379) | 3<br>(0-12) | 56<br>(17-320) | 1<br>(0-24) | 375<br>(235-1287) | 3,922<br>(1,492-10,807) | 1,681<br>(502-3,517) | 807<br>(324-2,718) | 1,434<br>(666-4,572) |  |
| Primary adrenal<br>insufficiency | 0-24h |  |  |  |  |  | 0-6h | 6-12h | 12-18h | 18-24h |
|  | C<sub>max</sub><br>(nmol/L) | T<sub>max</sub> (h) | C<sub>min</sub><br>(nmol/L) | T<sub>min</sub> (h) | ΔC<sub>max</sub>-C<sub>min</sub> | AUC<br>(nmol*h/L) | AUC<br>(nmol*h/L) | AUC<br>(nmol*h/L) | AUC<br>(nmol*h/L) | AUC<br>(nmol*h/L) |
| Patients with primary adrenal insufficiency (N=10) receiving 200mg hydrocortisone over 24 hours in four different delivery modes |  |  |  |  |  |  |  |  |  |  |
| ORAL<br>(50mg/6h) | 1423<br>(1083-2457) | 0.5<br>(0.5-1.5) | 277<br>(64-398) | 6<br>(0.5-6) | 1089<br>(834-2393) | 15,267<br>(8,591-22,417) | 3,807<br>(2,471-5,731) | 4,056<br>(1,839-6,348) | 4,200<br>(2,539-5,700) | 3,944<br>(2,208-5,600) |
| IM<br>(50mg/6h) | 1152<br>(830-1345) | 1<br>(0.5-2) | 289<br>(148-453) | 6<br>(6-6) | 844<br>(581-1151) | 14,950<br>(10,383-20,102) | 3,887<br>(2,864-5,200) | 4,055<br>(2,429-5,296) | 3,781<br>(2,789-5,135) | 3,866<br>(2,711-5,305) |
| IVI<br>(50mg/6h) | 1449<br>(1072-2432) | 0.5<br>(0.5-6) | 171<br>(0-375) | 6<br>(6-6) | 1239<br>(954-2261) | 13,413<br>(9,412-20,220) | 3,577<br>(2,415-4,852) | 3,466<br>(2,623-4,815) | 3,453<br>(2,440-6,084) | 3,425<br>(2,310-5,253) |
| CIV<br>(200mg/24h) | 836<br>(661-1073) | 7<br>(2-18) | 520<br>(388-617) | 20<br>(12.5-23.0) | 329<br>(232-551) | 14,649<br>(10,934-19,082) | 3,582<br>(2,685-5,025) | 4,067<br>(2,938-5,112) | 4,004<br>(3,033-5,069) | 3,712<br>(2,796-4,766) |

The table reports pharmacokinetic parameters determined from circulating serum total cortisol concentrations observed in 22 patients undergoing elective surgery with general anesthesia (knife-to-skin = 0h), and after administration of 200mg hydrocortisone over 24 hours to 10 patients with primary adrenal insufficiency.
Hydrocortisone was administered either as six-hourly bolus injection (ORAL, IM, IVI) or by CIV. All data are presented as median (range); numbers for the three different hydrocortisone bolus administration modes represent averages of the observations made during the four consecutive 6-hour intervals, while CIV data refer to the time period 2-24h (steady state was achieved at 2h during CIV).
Abbreviations: CIV: continuous intravenous infusion; $C_{\max}$ : maximum serum total cortisol concentration observed; $C_{\min}$ : minimum serum total cortisol concentration observed; IM: intramuscular; IVI: intravenous injection; $T_{\max}$ : time when the maximum serum total cortisol concentrations ( $C_{\max}$ ) were observed; $T_{\min}$ : time when the minimum serum total cortisol concentrations ( $C_{\min}$ ) were observed.

After reaching C_max_, both serum cortisol and cortisone concentrations gradually decreased back to pre-surgical baseline levels in the patients with short duration surgery, while circulating glucocorticoid concentrations remained increased in the group with longer-lasting surgery (Suppl. Fig. 1). The serum cortisol/cortisone ratio followed a similar pattern, with no difference between the two groups after 24 hours (Suppl. Fig. 1).

### Pharmacokinetics of stress dose hydrocortisone in patients with primary adrenal insufficiency

After administration of bolus hydrocortisone, C_max_ were achieved after a median time of 30 min (ORAL and IVI) or 60 min (IM), followed by a decrease to minimum concentrations (C_min_) after a median time of 360 min, i.e. before the administration of the next 6-hourly dose (Figure 3a-c and Table 2). By contrast, CIV administration of hydrocortisone led to serum cortisol concentrations persistently within the same range from around 2 hours after the commencement of infusion, without distinct peak and trough concentrations after achievement of steady state (Figure 3d). Serum cortisone concentrations remained stable throughout, with no notable differences between the four hydrocortisone delivery modes (Suppl. Fig. 2).

**Figure 3.**
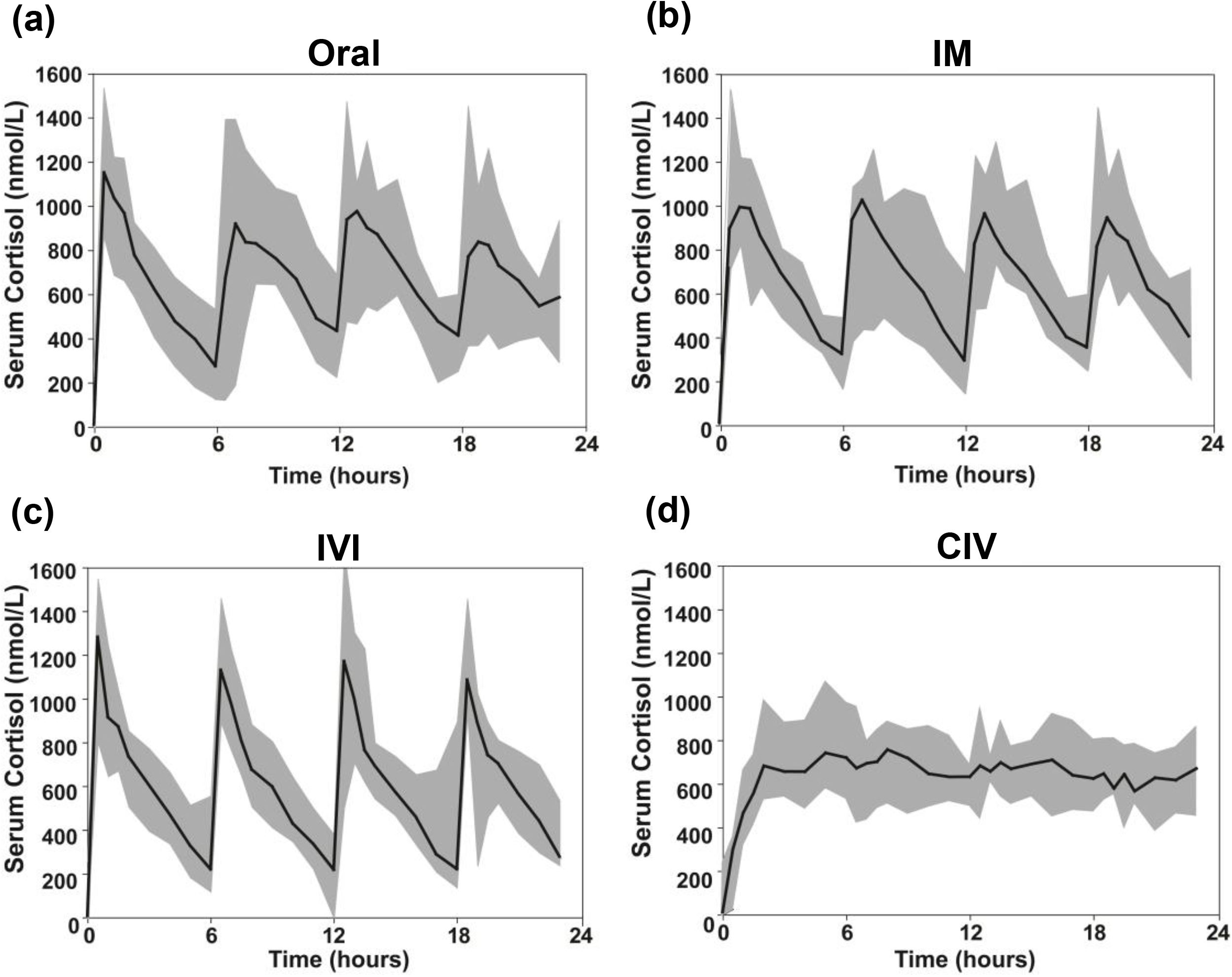
Serum total cortisol following hydrocortisone administration. Serum total cortisol (nmol/L) in 10 patients with adrenal insufficiency after hydrocortisone administered orally (ORAL), intramuscularly (IM), as intravenous boluses (IVI), and as continuous intravenous infusion (CIV). Data are presented as median (black line) and range (shaded grey area).

For all four hydrocortisone administration regimens, serum free cortisol concentrations at T_max_ (i.e. when C_max_ were observed) were significantly higher than those observed in patients exposed to different stress conditions, except for sepsis, where free cortisol tended to be higher (Suppl. Fig. 3a). Free cortisol during CIV at T_min_ (i.e. when C_min_ were observed) was significantly higher than in surgical patients at KTS and 4 hours into surgery, and after acute trauma, but significantly lower than in sepsis (Suppl. Fig. 3a). Free cortisol concentrations at T_min_ of the other hydrocortisone administration protocols were significantly lower than in sepsis, but did not differ from those observed during other stress conditions.

The pattern of 24-hour urinary glucocorticoid metabolite excretion was similar in patients receiving hydrocortisone in the IM, IV, and CIV administration modes while after oral hydrocortisone administration urine cortisol excretion was lower but cortisol metabolite excretion was higher (Suppl. Fig. 3b), indicative of a first-pass effect with rapid metabolism of cortisol to downstream tetrahydro-metabolites in the liver. Glucocorticoid metabolite excretion after exogenous hydrocortisone administration resembled the pattern observed in major trauma while patients with elective surgery and unstressed controls had a much higher proportion of cortisone metabolites (Suppl. Fig. 3b).

### Serum cortisol after hydrocortisone administration vs. serum cortisol during elective surgery

Serum cortisol concentrations observed in the 10 patients with primary adrenal insufficiency after hydrocortisone administration were plotted against the cortisol response of patients undergoing surgery of longer (N=11; Figure 4a-d) and shorter duration (N=11; Figure 4e-h). Initial peak cortisol concentrations after hydrocortisone administration in the primary adrenal insufficiency patients exceeded the concentrations observed during elective surgery in patients with preserved adrenal function. However, median cortisol concentrations after ORAL, IM, and IVI hydrocortisone administration decreased to trough levels below the median observed in patients undergoing longer-lasting surgery several hours before the scheduled repeat administration of bolus hydrocortisone (Figure 4a-c). By contrast, CIV hydrocortisone administration persistently maintained serum cortisol concentrations above the median of concentrations observed in patients undergoing elective surgery (Figure 4d).

**Figure 4.**
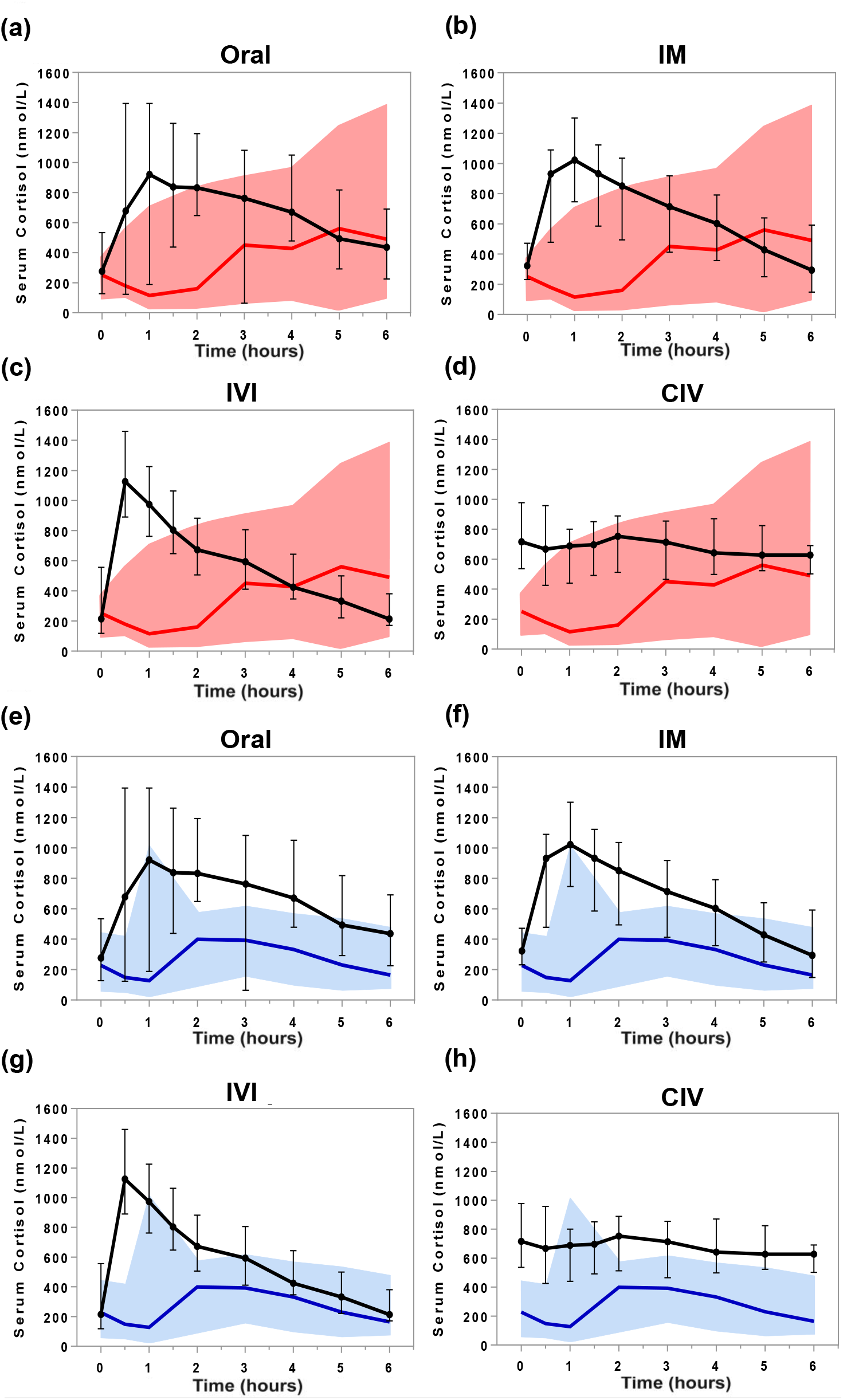
Comparison of serum total cortisol during elective surgery of longer duration and following hydrocortisone administration. Serum total cortisol concentrations (nmol/L) in 10 patients with adrenal insufficiency after the administration of 50mg hydrocortisone over 6 hours (black line: median, whiskers: range) in four different modes (orally [ORAL], via intramuscular [IM] or intravenous bolus injection [IVI], or as continuous intravenous infusion [CIV]) projected onto serum cortisol concentrations observed in patients undergoing elective surgery (Panels a-d, serum cortisol in 11 patients undergoing elective surgery of longer duration [red line: median; red shaded area: range]; Panels e-h, serum cortisol in 11 patients undergoing elective surgery of shorter duration [blue line: median; blue shaded area: range]) from time point knife-to-skin (KTS; 0 hours) to 6 hours post-KTS. All measurements were carried out by tandem mass spectrometry.

Serum cortisone concentrations in primary adrenal insufficiency and surgical patients showed a similar pattern; again, only CIV hydrocortisone administration achieved concentrations consistently above those observed in subjects undergoing elective surgery (Suppl. Fig. 4).

### Linear pharmacokinetic modelling of stress dose hydrocortisone administration

Next, we used the pharmacokinetic data obtained in the primary adrenal insufficiency patients undergoing exogenous hydrocortisone administration to model the most appropriate dose and mode of hydrocortisone delivery for raising cortisol concentrations quickly and sustain concentrations within the desired range, defined as above the median observed during elective longer-lasting surgery. Fitting to IVI, serum total cortisol concentrations yielded parameter estimates for the fixed effect (average) of initial response *Q* = 1,347nmol/L (SE 70nmol/L) and clearance rate *k* = 0.27 h^-1^ (SE 0.016 h^-1^). Random effect variances were calculated as (158nmol/L)^2^ and approximately 0, respectively.

Figure 5a depicts the 5^th^ and 95^th^ percentile range modelled on the serum cortisol concentrations observed after IV bolus injection of 50mg hydrocortisone dose; Figure 5b shows the predicted 24-hour serum cortisol concentrations. The model and fitted parameters were used to predict the serum cortisol responses to three alternative modes: 100mg hydrocortisone IV bolus injection (Figure 5c-d); initial 50mg (Figure 5e) and initial 100mg (Figure 5f) IV bolus injections, both followed by CIV infusion of 200mg/24h hydrocortisone. Modelling of these two regimens predicted that both would achieve the serum cortisol concentration range observed for longer-lasting elective surgery, with a near-instantaneous initial increase in serum cortisol concentration.

**Figure 5.**
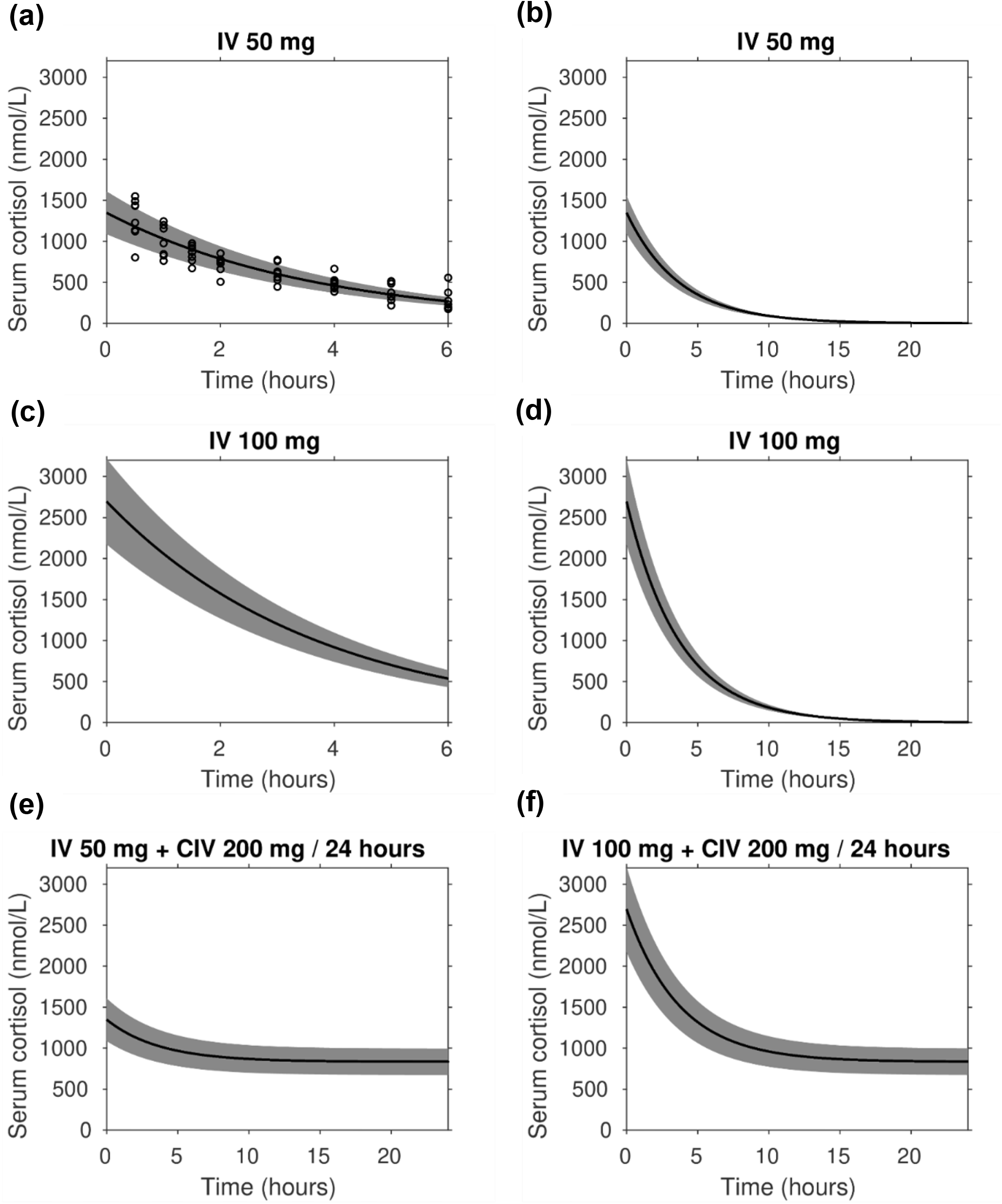
Linear pharmacokinetic modelling of stress dose hydrocortisone administration. Mixed effects linear pharmacokinetic modelling of serum cortisol in response to intravenous hydrocortisone administration modes. Serum cortisol concentrations are presented in nmol/L; the black lines show the fixed effect (central tendency) kinetics, the shaded gray area indicates 90% of between-patient variability. Panels a and b show cortisol measurements (circles) following 50mg IV bolus injection and the fitted model over 6 and 24 hours, respectively. The pharmacokinetic modelling was also used to predict the serum cortisol response to 100mg IV bolus injection over 6 and 24 hours (panels c and d, respectively), as well as initial 50mg (Panel e) and 100mg (Panel f) IV bolus injections followed by CIV infusion of 200mg/24h hydrocortisone.

## DISCUSSION

Patients with adrenal insufficiency are unable to mount a cortisol response to counteract a stressful event and, therefore, their regular replacement dose needs to be increased during major stress to avoid adrenal crisis (15). Nevertheless, no consensus exists regarding the optimal dose and hydrocortisone delivery mode during major stress, and current recommendations are empirical rather than evidence-based (7,16). This study is the first systematic dose-response study comparing the cortisol dynamics after administration of stress doses of hydrocortisone in patients with adrenal insufficiency to the acute cortisol response induced by surgery and other conditions of major stress. Our aim was to define the most clinically appropriate but still practically feasible regimen of hydrocortisone administration during major stress in patients with adrenal insufficiency, based on state-of-the-art tandem mass spectrometry measurements of circulating glucocorticoids. We found that continuous intravenous hydrocortisone was the only delivery mode that steadily maintained circulating cortisol in the range observed during major stress, while intermittent bolus administration of hydrocortisone resulted in frequent troughs with lower concentrations, thereby potentially exposing patients with adrenal insufficiency to periods of under-replacement, and hence the possibility of adrenal crisis, a life-threatening complication of cortisol deficiency.

In line with previously reported findings (17-19), we documented that serum total cortisol concentrations in all examined conditions of psychological and physical stress were increased above those observed in healthy, unstressed controls. The only exception was major trauma, with relatively lower total serum cortisol concentrations but increased serum free cortisol and 24-hour urinary cortisol, likely explained by the impact of blood loss in these patients.

Consistent with our previous systematic review and meta-analysis of the cortisol response to surgery (19), we observed an initial decrease in serum cortisol during elective surgery, which is likely to be linked to the induction of anesthesia. We observed higher cortisol concentrations during longer-lasting surgery, using reference standard tandem mass spectrometry for serum glucocorticoid analysis. This was also observed in a recent study in 93 patients undergoing elective surgery (20), with serum cortisol measurements carried out by immunoassay. In a previous meta-analysis of studies investigating the serum cortisol response to surgery (19) we did not find an impact of the duration of surgery on peri- and postoperative serum cortisol concentrations, likely explained by the heterogeneity of the studies included, which were also limited by the near exclusive use of immunoassays and lack of measurement of free cortisol. Linear pharmacokinetic modelling of stress dose hydrocortisone administration modes and doses combined with mixed effects regression identified continuous intravenous infusion of 200mg hydrocortisone over 24 hours as the most appropriate replacement regiment in patients with adrenal insufficiency exposed to major stress. Modelling indicated that this should be preceded by a one-off initial intravenous bolus of 50-100mg hydrocortisone to rapidly increase serum cortisol and shorten the time to steady state. We found that continuous intravenous hydrocortisone infusion was the only delivery mode to maintain cortisol concentrations persistently in the range observed during major stress including longer-lasting surgery. This regimen did not result in significant peaks and troughs in circulating cortisol, which were observed with the three hydrocortisone bolus administration modes (ORAL, IM, and IVI). Significant troughs potentially expose patients with adrenal insufficiency to under-replacement and the risk of life-threatening adrenal crisis, while supraphysiologic peaks might come with adverse side effects, as previously shown in the context of sepsis with an increased rate of hyperglycemic episodes (21).

A major strength of the present study is the use of reference standard tandem mass spectrometry for the measurement of circulating glucocorticoid concentrations, with all samples measured contemporaneously and with the same assay. Traditional immunoassays are associated with considerable inter-assay variation and potential cross-reactivity with other steroids, which may lead to over- and underestimations of true levels in critically ill patients with stress-induced stimulation of the hypothalamic-pituitary-adrenal axis (22,23). We also used mass spectrometry for the direct measurement of serum free cortisol, an important strength in comparison to studies who only employed indirect calculation of serum free cortisol utilizing cortisol-binding globulin, which is often inaccurate in the context of acute surgery and critical illness (19,24,25). Our previous systematic review and meta-analysis (19), only identified two studies measuring perioperative glucocorticoids by tandem mass spectrometry (26,27) and two studies directly measuring serum free cortisol in patients undergoing surgery (27,28).

One of the limitations of the present study is that, whilst serum cortisol was measured during elective surgery repeatedly over a 24-hour period, concentrations for the other stress conditions were measured at a single time point only. In the acute phase, sepsis causes a surge of circulating cortisol to persistently raised concentrations (17). Though we observed the highest serum cortisol concentrations in sepsis, it is unlikely that hydrocortisone doses higher than 200mg/24h would be required to cover patients with adrenal insufficiency in that situation, as critical illness results in a decrease in cortisol inactivation (29). Moreover, in the context of patients with sepsis but normal adrenal function prior to illness, an increase from hydrocortisone 200mg/24h to 300mg/24h did not impact on morbidity or mortality (30). In the present study, we did not assess the dynamics of cortisol metabolism during surgery and major stress. The previously reported reduced cortisol clearance during critical illness (29) may affect the requirements of hydrocortisone in patients with adrenal insufficiency and should be taken into consideration when interpreting our findings. A recent study reported that 100mg hydrocortisone/24h might be sufficient, though this was based on data collected in mostly secondary AI patients with likely residual cortisol biosynthetic capacity, with measurements carried out by immunoassays (31), Another limitation of our study is that the surgical group comprised mostly patients undergoing moderately invasive procedures. Thus, we cannot exclude that more invasive and longer-lasting surgeries could yield even higher serum cortisol concentrations. However, maximum cortisol concentrations were usually observed after the end of surgery in our patients, likely coinciding with withdrawal of general anesthesia and more invasive surgeries will be undertaken with the appropriately anesthesia and pain control regimens, thus not necessarily eliciting a higher cortisol response.

In conclusion, our data provide evidence that hydrocortisone stress dose cover during surgery, trauma, and major illness in patients with adrenal insufficiency should be provided by continuous intravenous infusion of 200mg hydrocortisone over 24 hours, following the administration of an initial intravenous hydrocortisone bolus of 50-100mg.

## Data Availability

The datasets generated during and/or analyzed during the current study are not publicly available but are available from the corresponding author on reasonable request.

## Notes

**Funding:** This work was supported by the Medical Research Council UK (program grant G0900567, to WA), the Oxfordshire Health Services Research Committee (NK), and the National Institute for Health Research (NIHR) Birmingham Biomedical Research Centre at the University Hospitals Birmingham NHS Foundation Trust and the University of Birmingham (grant reference number BRC-1215-2009, to WA and JML). AP is a Diabetes UK Sir George Alberti Research Training Fellow (grant reference number 18/0005782). IB is the recipient of a Robert and Elizabeth Strickland Career Development Award, the James A Ruppe Career Development Award in Endocrinology, and the Mayo Clinic Catalyst Award for Advancing in Academics.

### Competing Interest Statement

The authors have declared no competing interest.

### Funding Statement

This work was supported by the Medical Research Council UK (program grant G0900567, to WA), the Oxfordshire Health Services Research Committee (NK), and the National Institute for Health Research (NIHR) Birmingham Biomedical Research Centre at the University Hospitals Birmingham NHS Foundation Trust and the University of Birmingham (grant reference number BRC-1215-2009, to WA and JML). AP is a Diabetes UK Sir George Alberti Research Training Fellow (grant reference number 18/0005782). IB is the recipient of a Robert and Elizabeth Strickland Career Development Award, the James A Ruppe Career Development Award in Endocrinology, and the Mayo Clinic Catalyst Award for Advancing in Academics.
The views expressed are those of the authors and not necessarily those of the NIHR or the Department of Health and Social Care UK. The funders of the study had no role in the: design and conduct of the study; collection, management, analysis, and interpretation of the data; preparation, review, or approval of the manuscript; decision to submit the manuscript for publication.

